# Spiritual and religious aspects influence mental health and viral load: A quantitative study among young people living with HIV in Zimbabwe

**DOI:** 10.1101/2023.04.24.23289049

**Authors:** Ursula Wüthrich-Grossenbacher, Abigail Mutsinze, Ursula Wolf, Charles Chiedza Maponga, Nicholas Midzi, Masceline Jenipher Mutsaka-Makuvaza, Sonja Merten

## Abstract

**Introduction:** The role of religion and spirituality as social determinants of health have been widely discussed in and outside the World Health Organization. Studies among people living with Human Immunodeficiency Virus (HIV) describe positive and negative influences of religion and spirituality on health outcome. With a HIV prevalence of 14.8% for females and 8.6% for males, and 22’000 Aids related deaths in 2020, HIV infection remains a life-threatening condition in Zimbabwe, especially for young people. The aim of this research was to measure the influence of religion and spirituality on the health outcome of young people living with HIV in Zimbabwe.

**Methods:** A quantitative questionnaire with three different validated measures of religion and spirituality (Belief in Action Scale, Brief Religious Coping Index, Religious and Spiritual Struggles Scale), demographic, cultural, behavioral, and health questions was administered to 804 young Zvandiri program clients in rural, urban, and peri-urban Zimbabwe between July and October 2021. Regression analysis established significant relations between the result of the three different measures and mental health and viral load results.

**Results:** Religious coping significantly reduced the probability of common mental disorder, while high religious activity increased the risk. The Religious and Spiritual Struggles scale proved to be a reliable indicator of higher viral loads, risk for treatment failure, and the probability of common mental disorder.

**Conclusions:** The Religious and Spiritual Struggles scale should be used and validated in other sub-Saharan contexts. It could serve as a new diagnostic tool for the early detection and prevention of treatment failure as well as of common mental disorder.

## INTRODUCTION

In 1984 the 37^th^ World Health Assembly passed a resolution asking to include “The spiritual dimension in the Global Strategy for Health for All by the Year 2000”.(1) The delegates were convinced that spiritual wellbeing should be considered as the fourth dimension of wellbeing. The reflection on this resolution begins with the definition of the term spiritual: “a phenomenon that is not material in nature but belongs to the realm of ideas that have arisen in the minds of human beings, particularly ennobling ideas”.(2) The reflection states that philosophical, religious, moral or political ideologies influence the physical, mental and social wellbeing of people and that such ideologies and values also constituted the moral basis of the Global Strategy for Health for All. It points out that the target of a socially productive life has a non-material connotation and stresses the importance of people having an awareness of the different factors affecting their health and their health seeking behaviour.(2) Winiger and Peng-Keller recently conducted a research project on the ‘spiritual dimension’ of health as discussed in the World Health Organization (WHO). They suggest that the WHO should continue to interact with religious stakeholders.(3)

Increasingly epidemiologist recognize religion as a social determinant of health.(4) Hence, empirical studies about the influence of religion/spirituality (R/S) on health are gaining importance.(5) In 1999 the Fetzer institute published The Brief Multidimensional Measurement of Religiousness/Spirituality for Use in Health Research (BMMRS). The BMMRS assesses 12 key dimensions of R/S identified as relating to health outcomes: daily spiritual experiences, meaning, values, beliefs, forgiveness, private religious practices, religious/spiritual coping, religious support, religious/ spiritual history, commitment, organizational religiousness, and religious preference, and self-ranking of religiousness and spirituality.(6) Since then, many different R/S measurements have been developed that assess one or more of these dimensions. A growing number of studies, including those with study populations of people living with Human Immunodeficiency Virus (PLHIV), have shown that R/S can influence health in positive and negative ways. Kendrick published a systematic literature review of studies among PLHIV in the USA. Of the 33 empirical studies included, 24 studies found at least one measure of R/S associated with better adherence and clinical health outcomes, twelve studies found at least one measure of R/S associated with poorer adherence and clinical health outcomes, and seven studies found at least one measure of R/S to have no significant association with outcomes.(7) However, there was little consistency in regard to the measurement of R/S. In 2018 Doolittle et al. also published a systematic review of literature about the relationship between R/S and Human Immunodeficiency Virus (HIV) clinical outcomes. They found ten studies reporting a positive association of R/S with a measurable clinical outcome, two studies reported neutral associations and one study that identified aspects of R/S that had both negative and positive associations.(8) In our own literature review about the importance of considering religious and spiritual ontologies in the care of PLHIV in Zimbabwe, we concluded that R/S (including traditional practice) play an important role in the life of PLHIV in Zimbabwe, influencing the health, wellbeing, and access to care in positive and negative ways.(9) We therefore believe that R/S aspects and actors should be included in the comprehensive care of PLHIV.

Zimbabwe has a HIV prevalence of 14.8% for females and 8.6% for men (10). With 22’000 Aids related deaths in 2020 (11), HIV infection remains a life-threatening condition, especially for young people. Fortunately, with the development of highly active antiretroviral therapy, HIV virus suppression rates have increased. In our cohort of Zvandiri beneficiaries (12), viral load suppression has increased from 77% in 2018 to 88% in 2020 (13) (at that time Zimbabwean’s Ministry of Health and Child Care (MoHCC) defined suppression as below 1000 copies/ml). Fifteen years ago, Ironson and Kremer showed that an increase in religiousness/spirituality from before to after finding out that one was HIV-positive was significantly related to change in viral load results over four years.(14) Today, with improved treatment, can we still find an effect of R/S on the viral load of PLHIV? Have R/S become neglectable factors for young people living with HIV? The aim of this study was to measure the impact of R/S aspects on the clinical outcome (viral load and mental health status) of adolescents and young people (aged 14-24 years) living with HIV in Zimbabwe.

### Measuring religion

How can R/S be defined, or more so, be scientifically measured? Religious expression, practice and beliefs are dynamic. Definitions vary from organized to unorganized, from personal to group, from theistic to non-theistic, from intrinsic to extrinsic and so on. In this paper the term “religion” refers to organised and/or shared faith practice or belief and the term “spirituality” refers to the way people relate to the transcendent, including traditional practices. The participants of this study come from a highly religious society. The majority identifies with the Christian belief. However, in the Zimbabwean context, Christianity and traditional belief practices exist alongside and are sometimes practiced simultaneously. This is especially true when it comes to health issues. Shoko explains that in the Shona understanding most conceive and practice healing holistically, embracing not only the physical conditions, but also the spiritual, psycho-emotional, social, and ecological dimensions.(15)

Hill and Pargament suggest to use several measures to assess religious and spiritual aspects.(16) Thus, we decided to use three measures, that together cover most BMMRS key dimensions and have been widely used in different cultural and religious contexts. The Belief in Action Scale (BIAS) (17) measures the importance of religion in a person’s life. Doolittle found eight studies that explicitly evaluated religious involvement and spirituality with HIV viral load and Cluster of Differentiation 4 (CD4) cell count. Most of them showed a positive association between high religious involvement and clinical outcome.(8) However, in our own research we found that conforming to the teachings of some religious faith community can have a negative impact on health (e.g. banning of contraceptives, or Western medicine).(9) The Brief Religious Coping index (Brief RCOPE) (18) measures positive and negative religious coping. Most studies link positive religious coping with better health. E.g. Kremer et al who relate spiritual coping with a prediction of CD4-cell preservation and undetectable viral load over four years.(19) In a recent study from Brazil, resilience among PLHIV was significantly and positively correlated with positive religious coping and negatively correlated with negative religious coping.(20) In regard to R/S struggles, Wilt, Exline and Pargament recently pointed out that while more intense R/S struggles normally associate with increased and diverse efforts to cope, R/S strugglers may be hesitant to rely on God.(21) In other words, they might use secular techniques of coping or self-centered coping (corresponding to below negative coping item “trying to make sense of the situation without relying on God/spiritual force as partners”). The RSS (22) measures the prevalence of religious and spiritual struggles in a person’s life. In 2010 Trevino et al. showed that spiritual struggle was associated with a detectable viral load as well as more HIV related symptoms.(23)

## METHODS

This study is part of a larger mixed method study entitled “Impact of religion/spirituality (R/S) on HIV therapy in different health settings in rural and urban Zimbabwe with a special focus on adolescent and young people and the current Covid-19 pandemic” among Zvandiri’s cohort of adolescents living with HIV. The primary researcher is affiliated with the National Institute of Health Research and licenced by the Research Council of Zimbabwe. The study conforms to the principles embodied in the Declaration of Helsinki. Ethical approval was provided by the Medical Research Council Zimbabwe (MRCZ/A/2701). Informed consent was obtained from all adult participants and caregivers in case of minors. Minors assented to the study.

We used the STROBE cohortreporting guidelines.(24)

### Study population

The 804 participants (aged 14-24) for the quantitative study part were beneficiaries of Zvandiri’s peer support program in Zimbabwe (25). Participants were selected from MoHCC facilities in seven districts. The facilities were purposefully chosen to ensure a good balance of location (rural, urban, peri-urban), religious environment (main churches, traditional religions, Apostolic sects), languages and cultures (Shona, Ndebele), and facility structure (difference in user fees). Participants needed to have a current viral load result not older than 6 months.

#### Sample Size

Based on the literature review, we expected 20% of patients to have considerable/intense religious struggles. We further hypothized that patients with considerable/intense religious struggles have a 5-10% point difference in prevalence of viral non-suppression. At the time 85% of Zvandiri’s beneficiaries had an undetectable viral load according to MoHCC guidelines. Thus, calculating with

Alpha 0.05, Beta 0.2 and Power 0.8 and in consideration of missing viral load results because of the Covid-19, we aimed at 800 participants.(26)

### Questionnaire

The questionnaire collected following baseline data for the mother study: The R/S measures Belief in Action Scale (BIAS) (17), the Brief Religious Coping index (Brief RCOPE)(18), and the Religious and Spiritual Struggle Scale (RSS) (22), religious affiliation, beliefs regarding God, beliefs about spirits, opinions about the origin of HIV, types of healers and types of traditional medicines and practices involved in patients’ life, experience of violence, and other confounding variables and possible effect modifiers (demographic factors like age, location, education, civil status, income). The primary endpoint was the result of the mental health Shona Symptom Questionnaire (SSQ-14).(27) The secondary endpoint was the viral load result. Together with the entire questionnaire, the RSS was translated into the two major local languages Shona and Ndebele. The data collection team was recruited by Zvandiri among their community adolescent treatment supporters (CATS). After intensive training of all data collectors and a successful pilot in May, the quantitative data collection with the Open Data Kit (ODK) questionnaire in Shona, Ndebele, and English took place from July to October 2021. Due to the Covid-19 pandemic, most questionnaires had to be administered by phone, or were self-administered by an online public link. There was a daily check for answers indicating personal risk, with immediate referral of respective participants to the Zvandiri counsellor team. There was no loss to follow-up.

### Measures of Religion/Spirituality

BIAS: The basis for the content of the BIAS is the importance of religion in a person’s life. It inquires how individuals spend their time, talents, and financial resources, what individuals say is important in their life, and to what degree they conform to the religious teachings of their faith. The BIAS questions assess organizational and non-organizational religious activities, as well as degree of personal (intrinsic) devotion or commitment to one’s religious faith. The ten questions have answers on a 10-point Likert scale.

Brief RCOPE: We used the adapted version included in the BMMRS. Respondents are asked to think about how they try to understand and deal with major problems in their life. Three items assess positive religious coping, defined as “reflective of benevolent religious methods of understanding and dealing with life stressors”(28) and three items assess negative religious coping reflecting underlying spiritual tensions and struggles.(18) On a Likert scale from 1-4 participants mark to what extent each of the following is involved in the way they cope: I think about how my life is part of a larger spiritual force. I work together with God/spiritual force as partners. I look to God/spiritual force for strength, support, guidance. I feel that stressful situations are God’s (or spiritual force’s) way of punishing me for my sins or lack of spirituality. I wonder whether God/spiritual force has abandoned me. I try to make sense of the situation and decide what to do without relying on God/spiritual force.(6)

RSS: The twenty-six-item RSS measures six domains: divine (negative emotion centered on beliefs about God or a perceived relationship with God), demonic (concern that the devil or evil spirits are attacking an individual or causing negative events), interpersonal (concern about negative experiences with religious people or institutions; interpersonal conflict around religious issues), moral (wrestling with attempts to follow moral principles; worry or guilt about perceived offenses by the self), doubt (feeling troubled by doubts or questions about one’s r/s beliefs), and ultimate meaning (concern about not perceiving deep meaning in one’s life).(22) Participants answer on a Likert scale from 1-5.

#### Validity check

The three measures were checked for reliability and validity using Cronbach’s alpha. Construct validity was assessed using the Mokken Scale Analysis (29) and exploratory factor analysis for the RSS.(30)

### Analysis of survey data and primary and secondary outcome

The data was analyzed using the statistical software STATA version 17.0 (Stata Corp, College Station, TX, USA).

We present descriptive statistics showing the sociodemographic characteristics of participants. The primary outcome variable was the mental health status. Average ≥ 8 in the 14 items Shona Symptom Questionnaire (SSQ-14)(27) indicate that there is a probability of common mental disorder.

The secondary outcome was the current viral load result. The variables were defined according to the European Aids Clinical Society’s guidelines.(31)

Viral load: **1** ≤ 50 copies/ml, **2** >50 copies/ml<=200 copies/ml, **3** >200 copies/ml Target non-detectable (TND): **1** ≤ 50 copies/ml, **0** >50 copies/ml

Failure: **1** >200 copies/ml, **0** <200 copies/ml

We used multilevel mixed-effects ordered logistic regression (meologit) and multilevel mixed-effects logistic (melogit) regression to measure the association between viral load and mental health status results and the outcomes of BIAS, Brief RCOPE and RSS variables. Krause et al. suggest that socioeconomic status is associated with spiritual struggles. They tested following core hypotheses: “(1) individuals with lower levels of educational attainment are more likely to encounter chronic economic difficulties; (2) people who experience ongoing financial strain are more likely to live in rundown neighborhoods; (3) people who live in dilapidated neighborhoods will be more angry than their well-to-do counterparts; (4) people who are more angry will, in turn, be more likely to experience spiritual struggles; and (5) greater spiritual struggles will be associated with more symptoms of physical illness.”(32) Besides controlling for age, gender and education, we also controlled for location (= health-facility) in our regression. We did not control for income, as almost all our participants are poor.

### Patient and Public involvement

The content of the questionnaire was a result of discussions with local researchers and experts from the medical, theological, traditional, and social science field. The data collection team consisting of Zvandiri’s senior research coordinator and former and current CATS were involved in editing and testing the questionnaire. This included assessment of the possible psychological burden for data collectors and respondents, defining “red flag answers” and referral procedures, confidentiality issues, and logistical and financial constraints (connectivity, access to phones, re-imbursement of air-time costs) and the time required to participate in the research. During the analyzing process, CATS were asked to comment on the results. Did the results reflect the impression the CATS had while conducting the data collection? CATS are peers of the participants of this study.

## RESULTS

Only 2 participants had missing viral load results and were excluded. All other 802 participants had no missing variables and were included in the analysis.

Chronbach’s alpha and Mokken Scale Analysis were used to ensure the validity of the three measurements in our context with young PLHIV in Zimbabwe.

### Validity Check of BIAS, Brief RCOPE and RSS

#### BIAS

Mokken Analysis identified one scale with six items. 4 items were excluded: What has the highest priority in your life now/what is most important? To what extend have you decided to place your life under God’s direction? What percentage of your income do you give to your religious institution, faith group or to other R/S causes? To what extend have you decided to conform your life to the teachings of your religious faith? We excluded church offerings, as we think, this item is not relevant in our context of extreme poverty. We created a standardized variable ‘religious frequency’ with alpha of the six items identified by Mokken Analysis. Out of the six items in the ‘religious frequency’ variable, two items relate to public practice and 4 items to private practice. Cronbach’s alpha for the new standardized Frequency variable was 0.7. We kept Life Priority (binary: 10 = God, 1 = everything else), God_commitment (place life under God’s direction) and Religious_commitment (conform life to religious teaching) as separate variables.

#### Brief RCOPE

Cronbach’s alpha for all six items was 0.7. Mokken Analysis identified the two dimensions of the original scale: Positive Coping with three items and Negative Coping with three items.

#### RSS

Cronbach’s alpha of all 26 items was 0.9. Mokken analysis of the RSS resulted in four sub-groups different to the original. Thus, we conducted an exploratory factor analysis (EFA) which also resulted in four sub-groups, partly corresponding to the Mokken sub-groups. The EFA sub-groups were coherent, culturally relevant, and were used for further analysis by creating four new binary variables: Alienation, Confusion, Conflict/Doubt and Religious Zeal. A detailed explanation of the RSS validation and the resulting new sub-groups/variables is available in our previous publication.(30)

### Participants Sociodemographic

60% of the participants of this study were below 18 years and 40% were between 18-24 years of age. 61.3% were females, 38.4 males, and 0.3% chose “other” gender. 49% of participants were from an urban environment, 13% lived in peri-urban settings and 38% were from the rural area. 86% of adult participants were earning less than the equivalent of 25 US Dollars per month. 27% never went further than primary school (Grade 7). Less than 10% were married.

The majority of our participants belonged to a Christian church. Percentage of TND, treatment failure, and probability of common mental disorder differentiated according to religious association (Table 1). The role of religious affiliation as predictors of current viral load results has been described in a paper that is in review. (33)

**Table 1:**
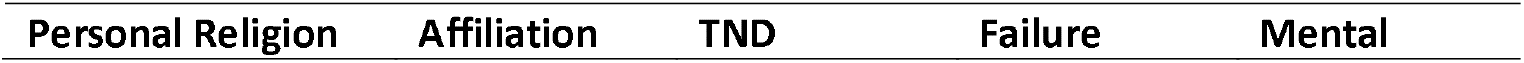
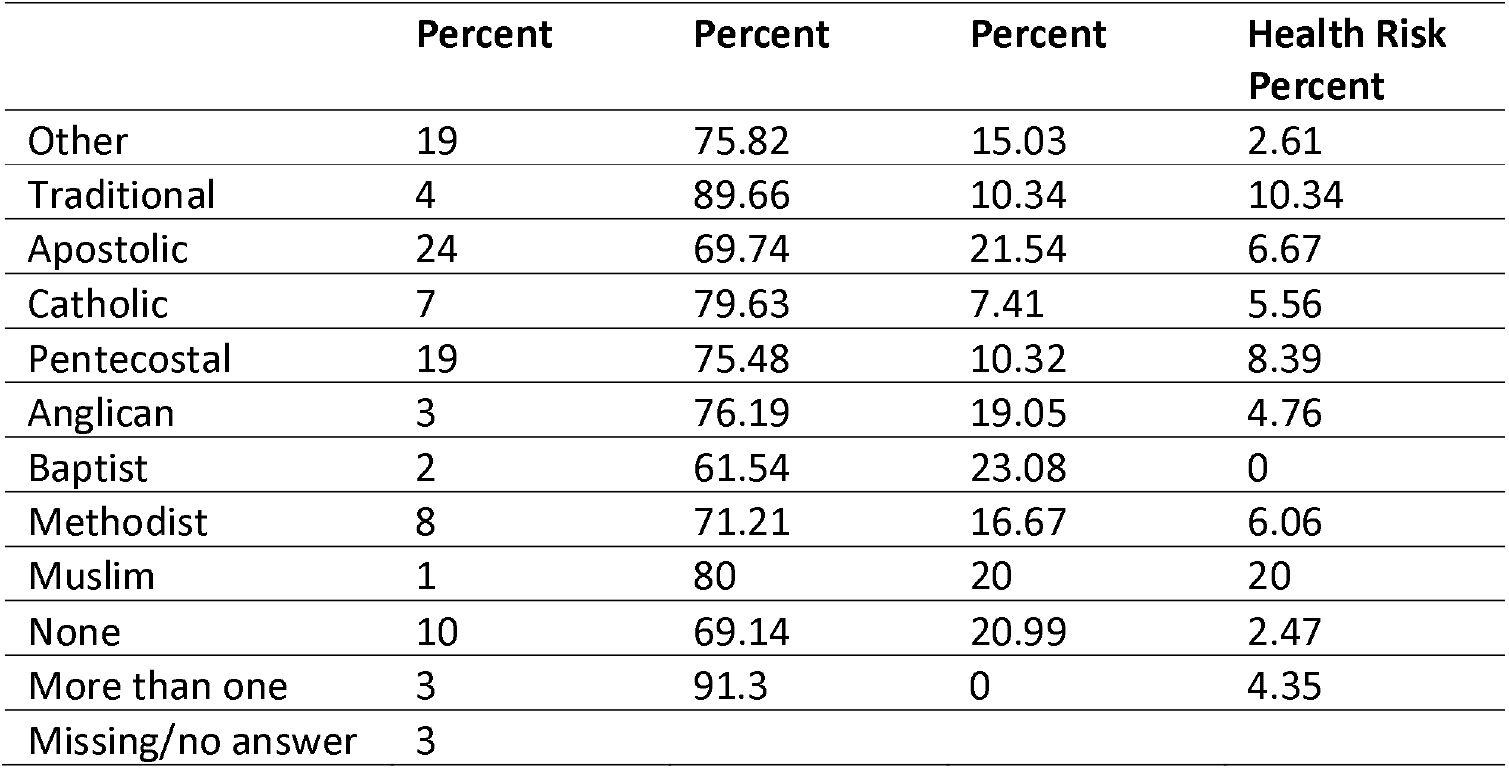
Participants Religious Affiliations (multiple answers were possible) and corresponding percentage of TND, treatment failure and probability of common mental disorder.

| Personal Religion | Affiliation | TND | Failure | Mental |
| --- | --- | --- | --- | --- |

|  | Percent | Percent | Percent | Health Risk<br>Percent |
| --- | --- | --- | --- | --- |
| Other | 19 | 75.82 | 15.03 | 2.61 |
| Traditional | 4 | 89.66 | 10.34 | 10.34 |
| Apostolic | 24 | 69.74 | 21.54 | 6.67 |
| Catholic | 7 | 79.63 | 7.41 | 5.56 |
| Pentecostal | 19 | 75.48 | 10.32 | 8.39 |
| Anglican | 3 | 76.19 | 19.05 | 4.76 |
| Baptist | 2 | 61.54 | 23.08 | 0 |
| Methodist | 8 | 71.21 | 16.67 | 6.06 |
| Muslim | 1 | 80 | 20 | 20 |
| None | 10 | 69.14 | 20.99 | 2.47 |
| More than one | 3 | 91.3 | 0 | 4.35 |
| Missing/no answer | 3 |  |  |  |

### Participants’ health parameters (primary and secondary outcome variables)

Mental Health was checked with the SSQ-14. Average of more than 8 in the SSQ-14 indicates a probability of common mental disorder. 9.2% of our participants had an average of more than 8 in the SSQ-14. However, this was not significantly related to viral load results.

The viral load result was described with three different variables. Viral load for overall results with three categories, TND for undetectable viral loads, and Failure for viral load results indicating treatment failure (Table 2).

**Table 2:** Viral Load Results.

| Variable | Value | Result in Percent |
| --- | --- | --- |
| Viral load (overall) | ≤ 50 copies/ml | 74% |
|  | >50 copies/ml ≤ 200 copies/ml | 10% |
|  | >200 copies/ml | 16% |
| TND | ≤ 50 copies/ml | 74% |
| Failure | >200 copies/ml | 16% |

### Participants’ BIAS, Brief RCOPE, RSS results

#### Bias

*Frequency standardized (std):* Participants were asked: “On average, how much time do you spend for a religious activity?” Alpha of six items with Likert 1-10 standardized (std): Mean: -5.54e-09 / Standard Deviation (SD): 0.67

*Life Priority, binary:* 21% had the relationship with God as priority in their life.

God commitment: Participants were asked: “To what extent have you decided to place your life under God’s direction?” (Likert 1-10, one being lowest): Mean: 2.2 / SD: 3

*Religious Commitment:* Participants were asked: “To what extent have you decided to conform your life to the teachings of your religious faith?” (Likert 1-10, one being lowest): Mean: 7.7 / SD 2.4

Correlations between Bias variables: God commitment and religious commitment were moderately negatively correlated (0.5). Other correlations between the different variables were below 0.2.

#### Brief RCOPE

*Positive Coping*, 3 items (Likert Scale 1 A great deal, 2 Quite a bit, 3 Somewhat 4 not at all). Mean: 2 /SD 0.8

*Negative Coping*, 3 items (Likert Scale 1 A great deal, 2 Quite a bit, 3 Somewhat 4 not at all). Mean: 2.7 / SD 0.9

Participants were further asked: “To what extent is your religion/belief involved in understanding or dealing with stressful situations in any way?” (Likert Scale 1 very involved, 2 Somewhat involved, 3 Not very involved, 4 Not involved). Mean: 2.3 / SD 1

#### RSS

The 26 questions of the RSS have a five-point Likert Scale: 1: not at all/does not apply; 2: a little bit; 3: somewhat; 4: quite a bit; and 5: a great deal. Mean: 2.2 / SD 0.8. The cutting point for the experience of considerable/intense R/S struggles was a mean of 3. 16% of participants with average score ≥ 3 were considered to experience considerable/intense R/S struggles.

### Results of mixed-effects logistic regression BIAS and Brief RCOPE variables with results of viral load and mental health status

Choosing to have the relationship with God as priority in life was linked with a higher probability of common mental disorder but was not significantly related with viral load results (Table 3).

**Table 3:**
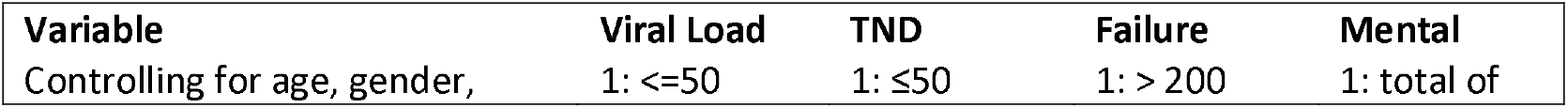
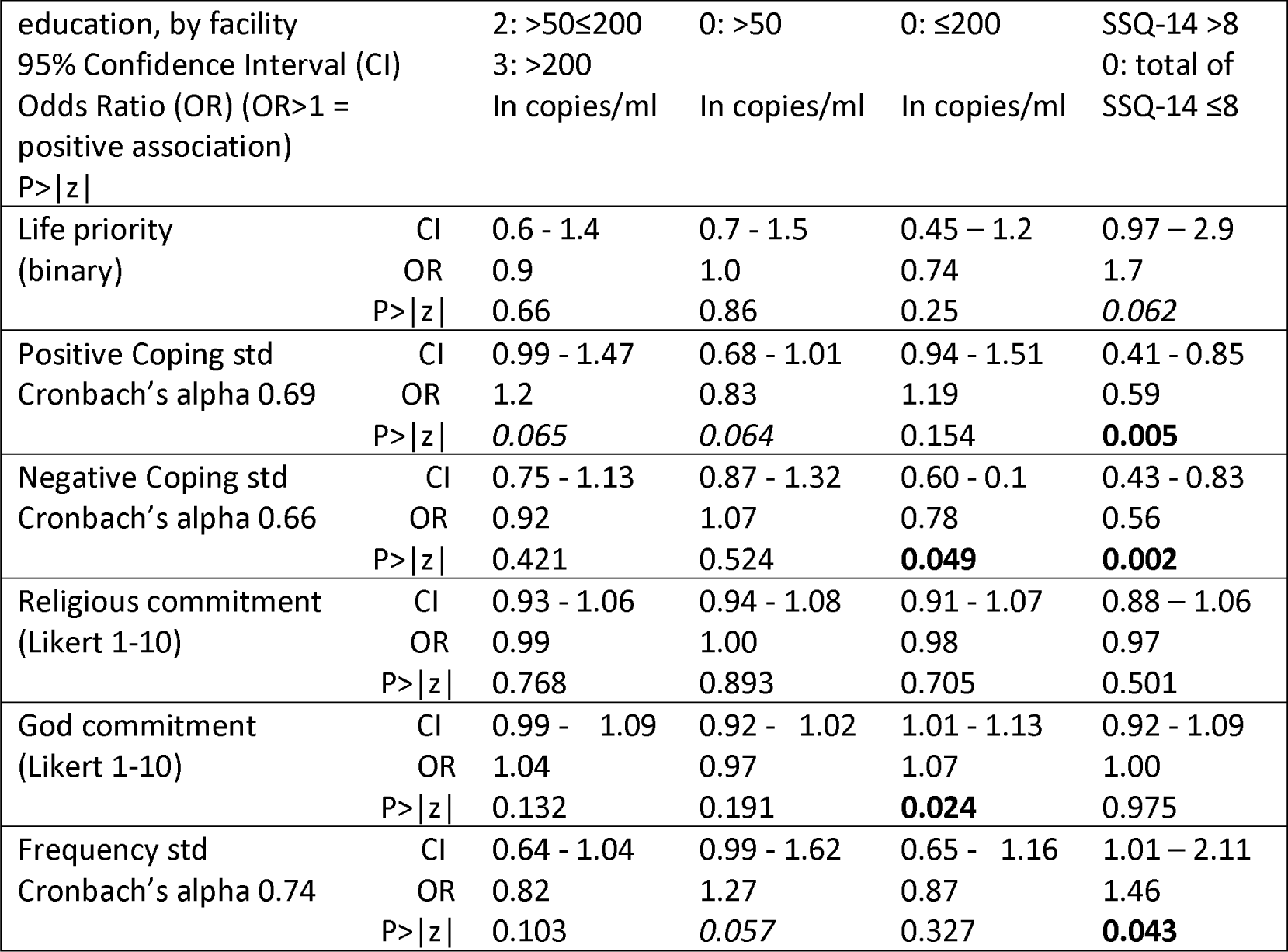
Individual Mixed-Effect Logistic Regression Results BIAS, Brief RCOPE, Viral Load, TND, Failure, and probability of common mental disorder. Significant relations = P>|z|. 0.05 are in bold, important relations = P>|z|0.05<0.1 in cursive.

| Variable | Viral Load | TND | Failure | Mental |
| --- | --- | --- | --- | --- |
| Controlling for age, gender, | 1: $\leq 50$ | 1: $\leq 50$ | 1: $> 200$ | 1: total of |

|  |  |  |  |  |  |
| --- | --- | --- | --- | --- | --- |
| education, by facility |  | 2: >50≤200 | 0: >50 | 0: ≤200 | SSQ-14 >8 |
| 95% Confidence Interval (CI) |  | 3: >200 |  |  | 0: total of |
| Odds Ratio (OR) (OR>1 = positive association) |  | In copies/ml | In copies/ml | In copies/ml | SSQ-14 ≤8 |
| P>\|z\| |  |  |  |  |  |
| Life priority (binary) | CI | 0.6 - 1.4 | 0.7 - 1.5 | 0.45 - 1.2 | 0.97 - 2.9 |
|  | OR | 0.9 | 1.0 | 0.74 | 1.7 |
|  | P>\|z\| | 0.66 | 0.86 | 0.25 | *0.062* |
| Positive Coping std Cronbach's alpha 0.69 | CI | 0.99 - 1.47 | 0.68 - 1.01 | 0.94 - 1.51 | 0.41 - 0.85 |
|  | OR | 1.2 | 0.83 | 1.19 | 0.59 |
|  | P>\|z\| | *0.065* | *0.064* | 0.154 | **0.005** |
| Negative Coping std Cronbach's alpha 0.66 | CI | 0.75 - 1.13 | 0.87 - 1.32 | 0.60 - 0.1 | 0.43 - 0.83 |
|  | OR | 0.92 | 1.07 | 0.78 | 0.56 |
|  | P>\|z\| | 0.421 | 0.524 | **0.049** | **0.002** |
| Religious commitment (Likert 1-10) | CI | 0.93 - 1.06 | 0.94 - 1.08 | 0.91 - 1.07 | 0.88 - 1.06 |
|  | OR | 0.99 | 1.00 | 0.98 | 0.97 |
|  | P>\|z\| | 0.768 | 0.893 | 0.705 | 0.501 |
| God commitment (Likert 1-10) | CI | 0.99 - 1.09 | 0.92 - 1.02 | 1.01 - 1.13 | 0.92 - 1.09 |
|  | OR | 1.04 | 0.97 | 1.07 | 1.00 |
|  | P>\|z\| | 0.132 | 0.191 | **0.024** | 0.975 |
| Frequency std Cronbach's alpha 0.74 | CI | 0.64 - 1.04 | 0.99 - 1.62 | 0.65 - 1.16 | 1.01 - 2.11 |
|  | OR | 0.82 | 1.27 | 0.87 | 1.46 |
|  | P>\|z\| | 0.103 | *0.057* | 0.327 | **0.043** |

Positive religious coping showed a significant association with lower probability of common mental disorder. However, it was also linked with higher viral loads and less TNDs (Table 3).

Negative religious coping was significantly related to lower probability of common mental disorder and less treatment failure (Table 3). To better understand this relationship, we conducted the same mixed-effects logistic regression with each of the three variables separately. Feeling that stressful situations are God’s (or spiritual force’s) way of punishing for sins or lack of spirituality was significantly linked to less treatment failure (OR: 0.83, CI: 0.7-0.98, P>|z|: 0.03) and lower probability of common mental disorder (OR: 0.7, CI: 0.5-0.8, P>|z|: 0.0). Wondering whether God/spiritual force had abandoned the participant was significantly linked to lower probability of common mental disorder (OR: 0.69, CI: 0.6-0.87, P>|z|: 0.001). Trying to make sense of the situation and deciding what to do without relying on God/spiritual force did not have any significant relation with viral load results or mental health status.

The degree of conforming to the teaching of one’s faith was not related to viral load results or mental health. However, claiming high commitment to place one’s life under God’s direction was significantly linked to increased treatment failure (Table 3).

More time spent on private and public R/S activities showed to be linked to a higher chance of TND and it was significantly associated with a higher probability of common mental disorder (Table 3).

In conclusion it can be said that lower probability of common mental disorder was significantly linked to both, positive and negative religious coping, while increased time spent on religious activities significantly increased the probability. The risk of treatment failure was significantly reduced with negative religious coping and increased with claiming high commitment to place one’s life under God’s direction (Table 3).

### Results of mixed-effects logistic regression RSS and RSS subdomains with results of viral load and mental health status

Considerable/intense RS struggles had a significant negative influence on viral load results and mental health. Looking at the different sub-domains of R/S struggles we found slight variations of the impact on viral load and mental health (Table 4).

**Table 4:**
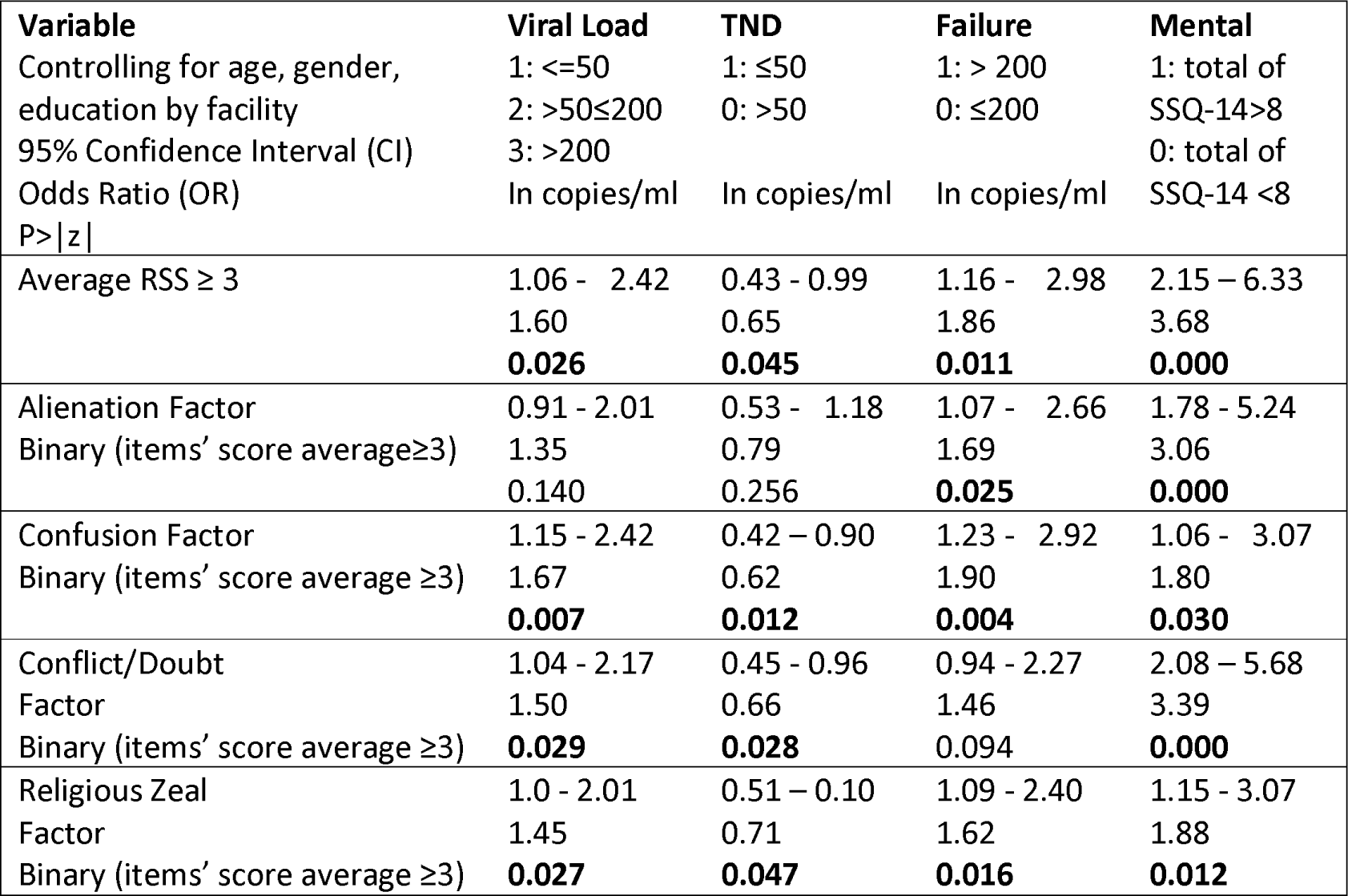
Individual Mixed-Effects Logistic Regression Results RSS and RSS subdomains, Viral Load, and probability of common mental disorder. Significant relations = P>|z|. 0.05 are in bold

#### Alienation Factor

“The RSS items corresponding to this sub-domain reflect feelings of hurt, isolation, misunderstanding and not being happy with the way religious activities are carried out.”(30) In a highly religious environment, low religiosity (lowest church affiliation, lowest God priority) may lead to pressure from family and society. This factor significantly related to higher probability of common mental disorder and higher risk of treatment failure. However, the Odds Ratio (OR) for risk of treatment failure was smaller than the OR of the overall RSS (Table 4).

#### Confusion Factor

The RSS items under this sub-domain express feelings of insecurity, loss of meaning, rejection, and anger. People in this sub-category had the highest percentage of multiple religious affiliations and conflicting beliefs about HIV. Most importantly they had the highest percentage of having stopped anti-retroviral treatment for religious reasons.(30) Hence, this factor was significantly associated with higher viral load results. It had the highest OR for high overall viral load and risk for treatment failure. It was also significantly related to higher probability of common mental disorder (Table 4).

#### Conflict/Doubt Factor

This sub-domain describes intellectual engagement with and questioning of religious, traditional, and spiritual norms, attitudes, and behaviors. In “a context of religious conformity, questioning and doubting may lead to the experience described by the RSS items belonging to this sub-domain. They refer to existential questions, struggle with the numinous, and a sense of not belonging.”(30) This was reflected in the highest OR of a significant higher probability of common mental disorder. The conflict/doubt factor was also significantly related to increased viral load and less full suppression (TND).

#### Religious Zeal

This sub-domain is linked to high religiosity, strong conviction in the existence of God and highest percentage of believing in the power of spirits and in the use of herbal supplements. It also had the highest percentage in moralizing views of HIV. “The RSS items belonging to this sub-domain express the struggle related to the failure of living up to religious standards. They describe self-questioning and self-condemnation, feelings of strained relationships with others, God, and the spiritual world, and the loss of ultimate meaning.”(30) Religious zeal was significantly associated with higher viral load results, less TNDs, higher risk of treatment failure, and it also significantly related to higher probability of common mental disorder (Table 4).

## DISCUSSION

Participants of this study were all part of an HIV program. The introduction of Highly Active Antiretroviral Therapy has significantly reduced HIV viral load results. However, according to European Aids Clinical Society’s guidelines, 16% of our participants’ blood results indicated treatment failure. This shows that additional, new, and innovative measures of care and support are needed to help these young PLHIV to better comply with the treatment regime. The findings of this study confirm the significant role of R/S as a social determinant of health for young PLHIV in Zimbabwe. Thus, integrating R/S aspects into the care and support of young PLHIV is not only desirable, but could become a key element for better adherence and treatment success. The findings of this study provide a basis for better identification of risk factors leading to treatment failure and indicate which aspects of R/S either hinder or further the individual health outcome of young PLHIV.

The role of religious coping remains ambiguous. While positive religious coping was significantly related to lower probability of common mental disorder, it was also related to higher viral load results. Further studies are needed to find out whether this is a case of reversed causality. Is positive religious coping used to cope with poor health? Negative religious coping had a positive influence on mental health and viral load results. Out of the three items, only the items directly related to negative religious coping were significant. Maybe guilt feelings and/or feelings of being abandoned by God stir up higher efforts or motivation for treatment compliance? Further studies are needed to investigate how negative religious coping influences viral load results and mental health outcomes.

For time spent on religious activities, there was a significant higher probability of common mental disorder. Also choosing the relationship with God as priority in life was linked to higher probability of common mental disorder. This is contrary to the findings of most other studies, that link religiousness (often measured by church attendance and private religious practice) to better mental health.(34) This might partly be explained by the gendered and spiritualized ideas about blame, transmission, and treatment of HIV, illustrated by O’Brian and Broom.(35) Pfeifer explains that R/S beliefs, norms, and attitudes influence the way life events (= stress, illness) are interpreted. The event might be given a spiritual meaning, or in other words, be spiritualized. This in turn, influences the psychosomatic mind body interaction.(36) Our findings show variation in viral load results and mental health between different religious affiliations. Kawachi points out that some aspects of regular service attendance might be detrimental to health. Considering the different religious teachings in different faith communities, the health effects of religious involvement may depend on the practices, teaching/preaching that person is exposed to.(5) Thus, in a religious context of blame, shame and stigma, the spiritualizing of HIV will mainly be negative, potentially leading to poorer mental health. This is one explanation. But causal relations are not always clear. Another explanation could be that increased religiousness is a coping mechanism for poor mental health. As our ‘religious frequency’ variable included more items related to private religious practice than public religious practice, we suggest that private religious practice (like meditation, prayer, serving, getting religious input (books, radio, TV etc.) might be the way of coping for young PLHIV.

Another ambiguous finding was that a high degree of placing one’s life under God’s direction increased the risk of treatment failure. Again, this might be explained by harmful religious teaching (only rely on God and not medicine), or on the other hand, the desire to place one’s life under God’s direction could have originated in the belief that this would help to get out of a poor health condition.

The findings of this study show that considerable/intense R/S struggles seem to be an important and reliable indicator of poorer mental and physical health for young PLHIV among Zvandiri beneficiaries. While the SSQ-14 identified 9.2% of participants with a probability of common mental disorder, the RSS seemed to be more sensitive, identifying 16% of respondents with considerable/intense R/S struggles. Furthermore, in contrast to the SSQ-14 scores, the results of the RSS were significantly associated with viral load results. The four sub-categories of the RSS provided useful information and descriptions of R/S struggles that were potentially harmful. This information could now be used to develop more holistic and comprehensive measures of care for young PLHIV. The findings further underline the importance of increased collaboration between the traditional and religious stakeholders and medical professionals.

We acknowledge that this study has some limitations. The study was limited to young PLHIV in a program setting in Zimbabwe. Results cannot be generalized outside this context. We strongly recommend the validation and use of the RSS in other contexts, especially in sub-Saharan Africa. Due to the Covid-19 pandemic most questionnaires were administered by phone. It was difficult to ensure privacy for respondents as they were confined to their homes, and others might have been around listening. This might have influenced some of the responses and explain the relatively low results of the SSQ-14 and RSS.

## CONCLUSION

In conclusion we can say that our study confirms the significant role of R/S aspects as social determinants of health. Especially the potential of the RSS as risk indicator for probability of common mental disorder, higher viral load, and treatment failure should be considered in new avenues of prevention and care for young PLHIV.

## Data Availability

All data produced in the present study are available upon reasonable request to the authors

## Abbreviations

BIAS: Belief in Action Scale
BMMRS: Brief Multidimensional Measurement of Religiousness/Spirituality
BriefRCOPE: Brief Religious Coping Index
CATS: Community Adolescent Treatment Supporters
CD4: Cluster of Differentiation 4
CI: Confidence Interval
EFA: Exploratory Factor Analysis
HIV: Human Immunodeficiency Virus
Melogit: Multilevel Mixed-Effects Logistic
Meologit: Multilevel Mixed-Effects Ordered Logistic
MoHCC: Ministry of Health and Child Care
ODK: Open Data Kit
OR: Odds Ratio
PLHIV: People living with Human Immunodeficiency Virus
RSS: Religious and Spiritual Struggles Scale
R/S: Religion/Spirituality
SD: Standard Deviation
SSQ-14: Shona Symptom Questionnaire std Standardized
TND: Target non-detectable
WHO: World Health Organization

